# Incorporating Human Movement Data to Improve Epidemiological Estimates for 2019-nCoV

**DOI:** 10.1101/2020.02.07.20021071

**Authors:** Zhidong Cao, Qingpeng Zhang, Xin Lu, Dirk Pfeiffer, Lei Wang, Hongbing Song, Tao Pei, Zhongwei Jia, Daniel Dajun Zeng

## Abstract

Estimating the key epidemiological features of the novel coronavirus (2019-nCoV) epidemic proves to be challenging, given incompleteness and delays in early data reporting, in particular, the severe under-reporting bias in the epicenter, Wuhan, Hubei Province, China. As a result, the current literature reports widely varying estimates. We developed an alternative geo-stratified debiasing estimation framework by incorporating human mobility with case reporting data in three stratified zones, i.e., Wuhan, Hubei Province excluding Wuhan, and mainland China excluding Hubei. We estimated the latent infection ratio to be around 0.12% (18,556 people) and the basic reproduction number to be 3.24 in Wuhan before the city’s lockdown on January 23, 2020. The findings based on this debiasing framework have important implications to prioritization of control and prevention efforts.

**One Sentence Summary:** A geo-stratified debiasing approach incorporating human movement data was developed to improve modeling of the 2019-nCoV epidemic.

## Main Text

Originating from Wuhan, Hubei Province, China, the 2019-nCoV epidemic has spread across China and 23 other countries, as of February 2, 2020 (*1–3*). As part of the national effort to contain the outbreak, an emergency lockdown measure was implemented city-wide in Wuhan on January 23, 2020, one day before the Chinese New Year Eve. However, according to official account, at least five million people already travelled from Wuhan to other parts of China before the city’s lockdown, causing many 2019-nCoV cases outside of Wuhan.

Estimating the key epidemiological parameters of 2019-nCoV has become a research priority with significant clinical and policy relevance. However, due to the limited emerging understanding of the new virus and its transmission mechanisms, results are largely inconsistent across studies. For example, estimates of the basic reproductive number range from 2.2 to 3.1 based on data collected in the early stage of the epidemic (*1, 3–5*). Attempts were also made to estimate the size of the epidemic, and forecast the progression of the epidemic with rather different conclusions (*1, 4, 5*).

Lacking accurate accounting or sampling of the ground truth as for the number of latently infected and undiagnosed infectious (L&I) persons is the primary cause for the widespread discrepancy in these estimates and forecasts. In particular, for the 2019-nCoV, one fundamental challenge is the under-screening and under-reporting biases in the early stage of the epidemic in its epicenter, Wuhan, due to limited testing and treatment resources while facing a major outbreak with a sudden onset. Note that this challenge is not unique to 2019-nCoV; see related discussions for SARS and other coronavirus outbreaks (*6, 7*).

To tackle this major challenge, we developed a geo-stratified debiasing estimation framework based on the following observation. On the contrary to Wuhan, other places in China have ample screening and treatment resources in relative terms, and started various stringent surveillance programs after the Wuhan lockdown, to screen and monitor people who travelled from Wuhan to these places. As such, the chance of under-screening and underreporting is much lower outside Wuhan, especially outside Hubei Province. Our approach stratifies mainland China into three zones, i.e., Wuhan, Hubei Province excluding Wuhan, and mainland China excluding Hubei (Fig. 1). Incorporating data capturing the size of the population-level movement from Wuhan to the destinations in other zones with the official confirmed 2019-nCoV case counts in these destinations, we obtained a reliable estimate of the latent infection ratio, defined as the proportion of L&I persons among the population, in Wuhan before the lockdown. In turn, this estimate enabled us to derive the basic reproductive number, the number of L&I persons in Wuhan before and after the lockdown, and assess the progression of the epidemic nationally.

**Fig. 1.**
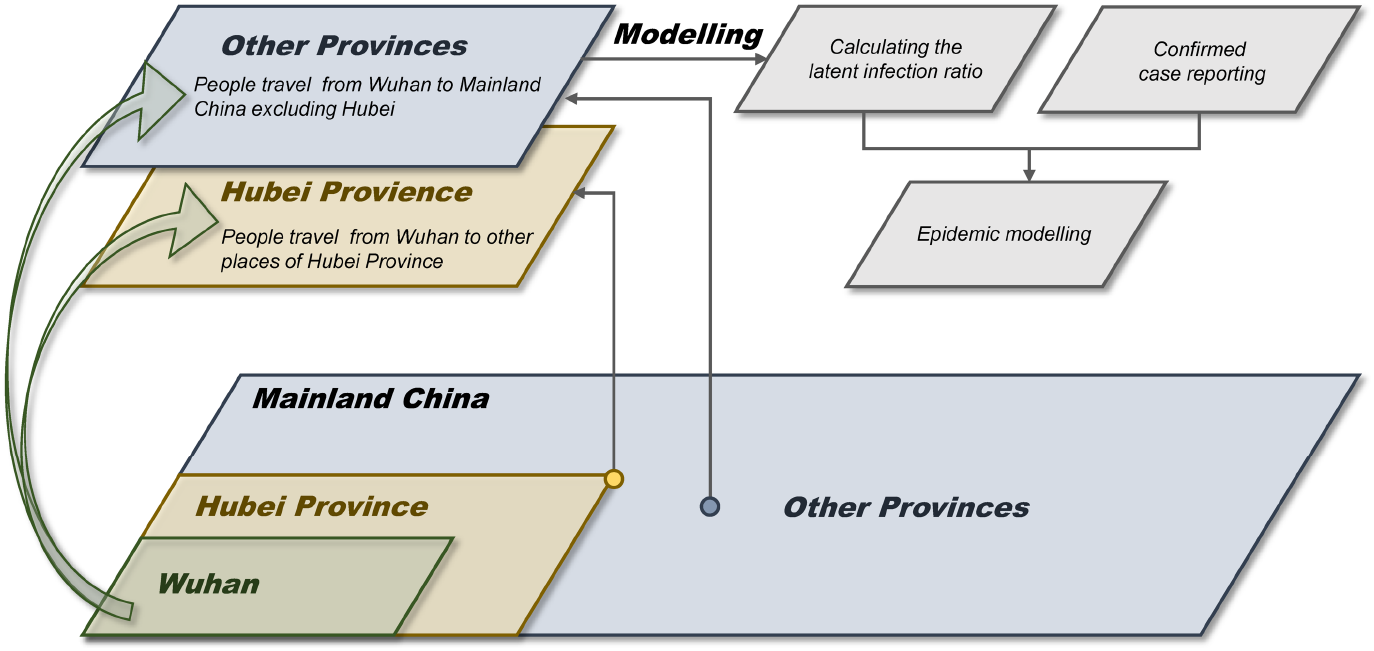
A geo-stratified debiasing estimation framework. The latent infection ratio was estimated from the number of people traveling from Wuhan to other destinations of mainland China. before the lockdown, and the confirmed 2019-nCoV case reporting data in these destinations. The estimation of the latent infection ratio enabled additional epidemiological modeling work such as calculating the 2019-nCoV’s basic reproduction number and inferring the actual size of the epidemic in Wuhan.

## Results

Fig. 2A shows the relationship between the number of people traveling from Wuhan during the period of January 16, 2020 to January 22, 2020, and the number of confirmed cases in 12 representative provinces and municipalities during the period of January 23, 2020 to January 29, 2020. The lag between these two time intervals was introduced to accommodate the commonly-assumed 7-day incubation period for 2019-nCoV (*3*). These representative provinces and municipalities are those that had received at least 30,000 travelers from Wuhan within a three-week window starting on January 1, 2020 and ending on January 22, 2020, prior to the lockdown.

**Fig. 2.**
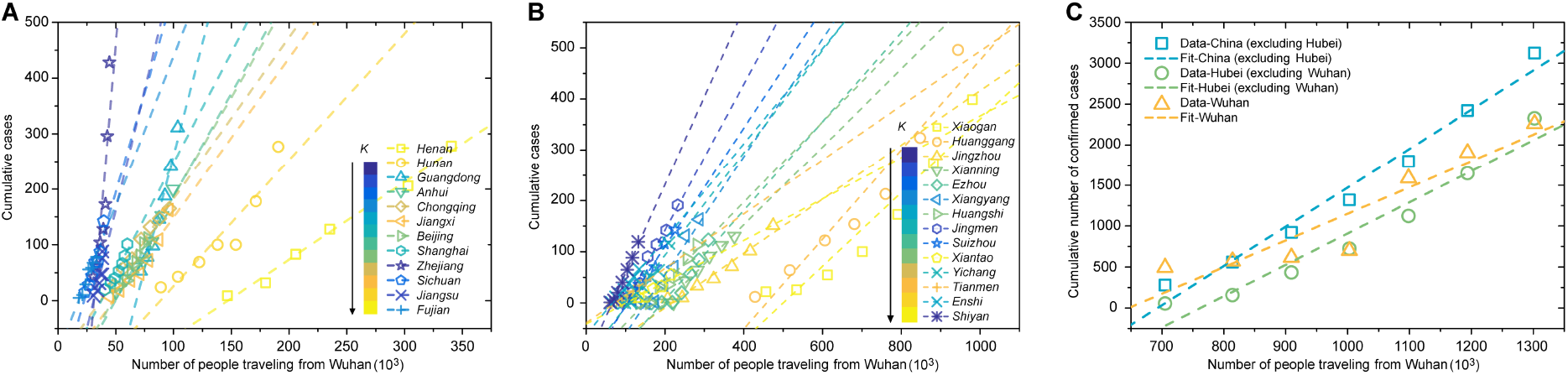
The relationship between the number of people traveling from Wuhan during the period of January 16, 2020 to January 22, 2020, and the number of confirmed 2019-nCoV cases during the period of January 23, 2020 to January 29, 2020. (**A**) For 12 provinces and municipalities each receiving 30,000+ travelers from Wuhan. (**B**) For 14 cities in Hubei each receiving 100,000+ travelers from Wuhan. (**C**) data aggregated for Wuhan, Hubei Province excluding Wuhan, and mainland China excluding Hubei, as well as linear fits.

Similarly, Fig. 2B presents the same relationship for 14 cities within Hubei Province. These cities had received at least 100,000 travelers from Wuhan. From Fig. 2A and Fig. 2B, we observe a statistically significant positive linear relationship between the number of travelers from Wuhan and the cumulative case count in most provinces and cities. For two provinces, Zhejiang and Jiangsu (in Fig. 2A), and two Hubei cities, Xiaogan and Huanggang (in Fig. 2B), we observe superlinear growth in the cumulative case counts, explainable by the known significant secondary local transmissions. Fig. 2C presents zone-level aggregated results.

Under our proposed geo-stratified debiasing framework, the latent infection ratio among the people traveling from Wuhan to other destinations in mainland China excluding Hubei within a three-week window prior to the lockdown, is estimated as 0.12% (95% CI: 0.09%-0.15%). Based on this latent infection ratio, we inferred that there were in total 18,556 (95% CI: 14,134-22,978) L&I persons in Wuhan before the lockdown, under the assumption that this latent infection ratio remained the same for people traveling from and remaining in Wuhan. Among these L&I persons, 10,887 (95% CI: 8,292-13,481) stayed in Wuhan on January 23, 2020, when the lockdown was initiated, and the remaining 7,669 (95% CI: 5,842-9,497) traveled to other parts of mainland China. Among the traveling L&I persons, 4,644-7,549 went to other places in Hubei Province, and 1,198 to 1,947 to places outside Hubei.

We followed a similar geo-stratified debiasing process, relying on more reliable case counts outside of Hubei to estimate the basic reproduction number *R*_*0*_ for 2019-nCoV. Our *R*_*0*_ estimate is 3.24, indicating that on average one infected case is expected to generate 3.24 secondary infected cases prior to isolation or death. This value is within the range of that of the 2003 SARS epidemic (2-5) (*8*), and higher than that of influenza (2-3) (*9*) and Ebola (1.5-2.5) (*10*). Compared to the WHO’s estimate (1.4-2.5) and other recently reported estimates (2.2-3.1) of 2019-nCoV (*1, 3–5*), our estimate is at the high end. This can be partially explained by the fact that most published estimates are based on all confirmed 2019-nCoV cases, including the Wuhan case counts, prone to the under-screening and underreporting biases. When considering the possibility of a large number of infected and infectious individuals who will not develop clinical disease, the actual value of *R*_*0*_ could be even higher.

Building on the estimated latent infection ratio, *R*_*0*_, and the number of L&I persons in Wuhan when the lockdown started, we developed a customized epidemiological model to simulate the progression of the 2019-nCoV epidemic after the lockdown of Wuhan (see supplementary materials for details). Given the stringent nature of the lockdown measures and the continuing underreporting bias in Wuhan, the characteristics of the epidemic within and outside Wuhan were treated differently.

Fig. 3A presents the model-predicted cumulative number of the confirmed 2019-nCoV cases for Wuhan, Hubei Province excluding Wuhan, and mainland China excluding Hubei. Fig. 3B shows the geographical distribution of estimated L&I persons on January 22, 2020, indicative of risks across mainland China.

**Fig. 3.**
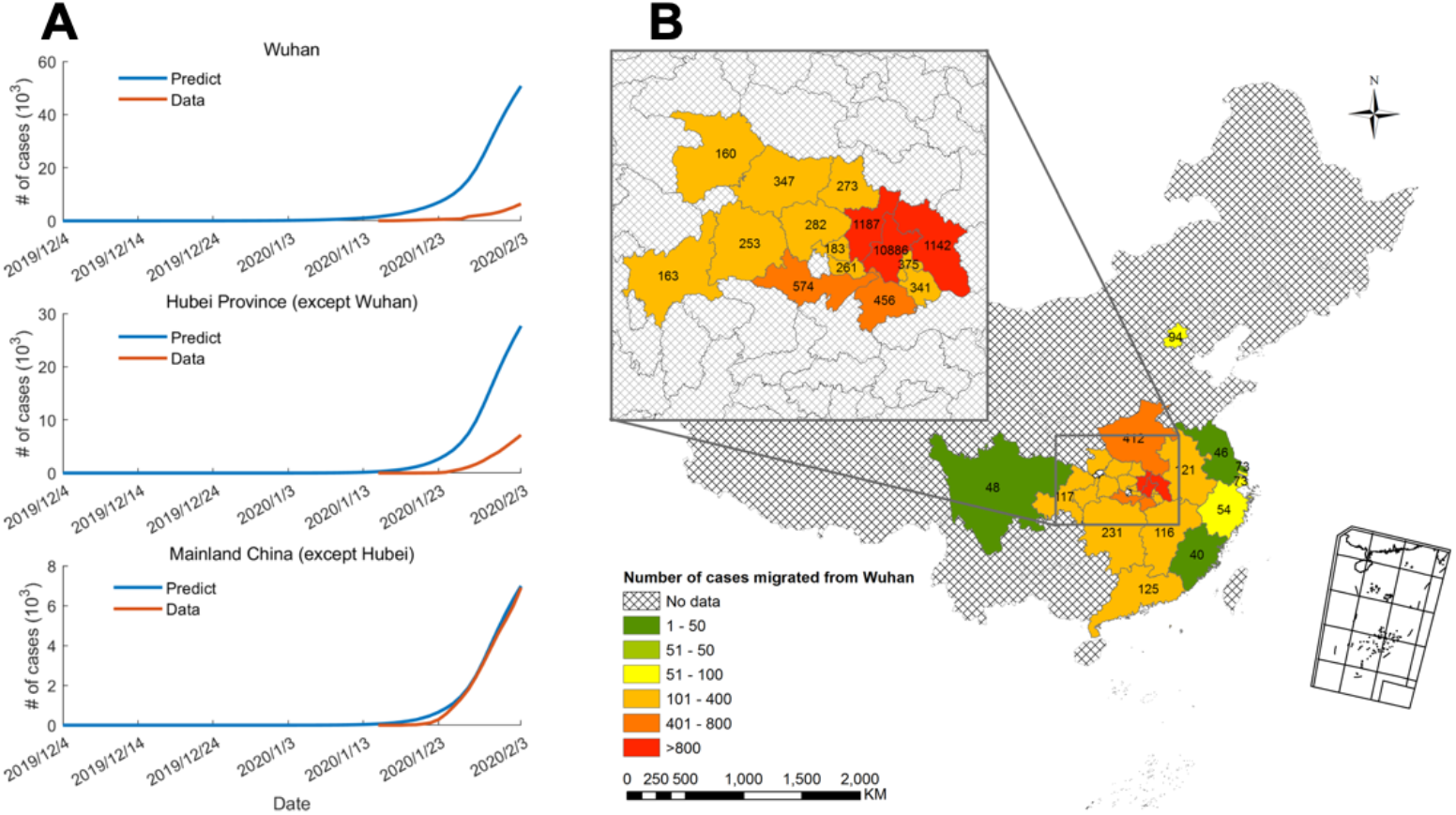
The model-predicted size of the 2019-nCoV epidemic. (**A**) The model-predicted cumulative number of confirmed cases in Wuhan, Hubei excluding Wuhan, and Mainland China excluding Hubei. (**B**) The number of 2019-nCoV L&I persons spread across Chinese Mainland on January 22, 2020.

## Discussion

This report introduces a geo-stratified approach for estimating the latent infection ratio in Wuhan with the presence of severe under-screening and underreporting biases. The estimated latent infection ratio enabled us to conduct further epidemiological investigations to better understand the ongoing 2019-nCoV epidemic.

Our findings have important implications with respect to the development of 2019-nCoV control and prevention policies in mainland China. First, the previous estimates of *R*_*0*_ are likely to have resulted in an underestimation of the transmissibility of 2019-nCoV, mainly due to the under-reporting bias during the early phase in Wuhan. Second, the actual epidemic size in Wuhan and Hubei is larger than what has so far been reported based on confirmed case data.

Third, for parts of Mainland China outside Hubei the risk of further outbreaks is likely to be limited, as long as locally occurring cases of secondary and tertiary transmissions can be detected and isolated effectively. The recent quarantine lockdown of Wenzhou was in response to detection of large numbers of secondary transmissions.

It is absolute key for being able to prevent this epidemic from becoming a pandemic that cases of clinical disease are detected and isolated as early as possible. In addition, all high-risk contacts need to be traced to identify secondary transmissions. The public health surveillance system in China is facing tremendous challenge give the population size and sensitivity of the system is to be enhanced. It is therefore paramount that available resources are used optimally. An understanding of the spatial distribution of the L&I persons is critical in this context, since they are the reservoir of clinical cases and of secondary transmission, although to a lesser extent than clinical cases. The annual migration around the Lunar New Year compounded the situation in that it immensely increased movements across the country. The human movement data utilized in this study provides a robust data source for estimating the likely dissemination of 2019-nCoV cases across the country (Fig. 3B), providing important epidemiological information that is much less affected by delayed- or under-reporting bias. The management of the epidemic over the coming weeks when people travel back from their hometowns to their work places will be critical for being able to bring it under control. We recommend that the national and local authorities process real-time human movement data through mathematical models to forecast the spatio-temporal dynamics of further 2019-nCoV outbreaks across Mainland China. This will allow developing early detection, isolation and quarantine strategies tailored to the very dynamic epidemiological situation, as well as identify potential problems in the implementation of local disease control policies and inform the design of any necessary adjustments.

## Data Availability

The human movement data is not available for sharing due to the constraint in the consent. All other data is available in the main text or the supplementary materials.

## Acknowledgments

The authors thank J. Yang, L. Li, H. Zhou, Y. Ye and K. Tang who helped with the initial data collection and cleaning.

## Author contributions

DDZ, ZC and QZ proposed the idea. ZC and QZ did the modeling and analytics. All authors contributed to the writing.

## Data and materials availability

The human movement data is not available for sharing due to the constraint in the consent. All other data is available in the main text or the supplementary materials. This study was approved by the Biomedical Research Ethics Review Board of Chinese Academy of Sciences Institute of Automation (approval #IA-202001).

## Supplementary Materials

### Materials and Methods

#### Daily counts of confirmed 2019-nCoV cases

The daily counts of confirmed 2019-nCoV cases were collected from the National Health Commission of the People’s Republic of China through its website accessible at http://www.nhc.gov.cn/xcs/yqtb/list_gzbd.shtml.

Additional information about these confirmed cases was retrieved from the websites of the official provincial and municipal health commissions throughout mainland China. A listing of these websites is provided by the National Health Commission of People’s Republic of China at http://www.nhc.gov.cn/xcs/yqfkdt/gzbd_index.shtml.

#### Human movement data

The human movement data used in the study consists of aggregated statistics provided by the two largest telecommunications operators in China, covering the daily counts of the number of mobile phone users traveling from Wuhan and reaching other destinations outside of Wuhan in mainland China, between January 1, 2020 and January 22, 2020. Given the very high mobile phone user penetration rate in China and the 80% market share held by these two operators, these statistics are expected to be representative of the actual size of the human migration originating from Wuhan. Note that some users may have returned to Wuhan before traveling outbound again. These users were counted more than once in the human movement dataset.

Due to the sensitive nature of the telecommunications data records, data processing and aggregation was conducted in-house in the telecommunications operators’ secure computing environment by their own staff, without any participation from the author team. The applicable law of “Provisions on Protection of Personal Information of Telecommunications and Internet Users (Mainland China)” (*11*), and the guidelines from the Global System for Mobile Communications Association (GSMA) on the protection of privacy in the use of mobile phone data, were followed (*12*). The authors have access to only exported data aggregated at the provincial and municipal level. No personal information was processed in the analysis of this study. This study was approved by the Biomedical Research Ethics Review Board of Chinese Academy of Sciences Institute of Automation (approval #IA-202001).

#### Estimation of latent infection ratio

Among all mainland China provinces and municipalities, we first chose those to which there were at least 30,000 people traveling from Wuhan between January 1, 2020 and January 22, 2020. From this selected list of 12 provinces and municipalities, Zhejiang Province and Jiangsu Province were excluded from our latent infection ratio estimation because of known substantial secondary local transmissions, given that the first objective of our study is to estimate the latent infection ratio in Wuhan during the early stage of the epidemic. For the remaining 10 provinces and municipalities, to accommodate the commonly-assumed 7-day incubation period for 2019-nCoV, we calculated the ratio between the count of confirmed cases between January 23, 2020 and January 29, 2020, and the number of people travelling from Wuhan during the period of January 16, 2020 and January 22, 2020. Table S1 presents these counts and the calculated ratios. Seven provinces and municipalities offered detailed information concerning secondary local transmissions when publishing daily case counts. Table S2 presents the cumulative data for these places.

The estimated latent infection ratio (0.12%) as reported in the main manuscript was calculated from the data reported in Tables S1 and S2, following the estimation model presented below.

Let *C*_*i*_ denote the number of cumulative confirmed cases in province *i*, and *N*_*i*_ the number of people traveling from Wuhan to province *i*. Mainland China has 31 provinces. Given the available data from the 10 selected provinces/municipalities, the latent infection ratio, the proportion of latently infected and undiagnosed infectious (L&I) persons in all people traveling from Wuhan, *l*, were calculated following the standard approach for ratio estimation (*13*), as follows.

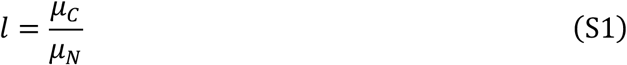

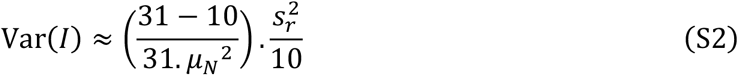

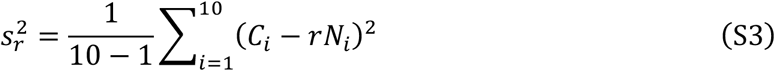

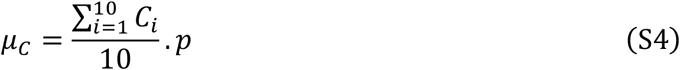

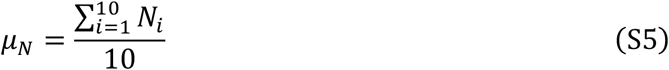

where *r* denotes the slope of the fitted linear relationship of (*N*_*i*_, *C*_*i*_), and p the average proportion of non-secondary cases (see Table S2), estimated around 0.776.

The resulting estimated latent infection ratio is 0.12% (95% CI: 0.09%-0.15%). Using this ratio, we in turn estimated that there were 18,556 (95% CI: 14,134-22,978) L&I persons (out of 15.34 million population) in Wuhan before the city’s lockdown. Among these L&I persons, 7,670 traveled to other destinations in Mainland China while the rest 10,886 remained in Wuhan.

#### The SEIRDC model

The standard SEIR model assumes four categories of persons: Susceptible, Exposed (corresponding to the latently infected cases in the main manuscript), Infectious, and Recovered. To investigate the 2019-nCoV epidemic, we adopted a the SEIRDC model (*14*), which enhances the standard SEIR model by introducing an additional category of persons, Dead, and an auxiliary variable *C*(*t*) that tracks the cumulative number of infectious persons during the outbreak. Furthermore, to take into account human movements, we customized the SEIRDC model for two epidemic areas: Wuhan and non-Wuhan (mainland China excluding Wuhan). The model is as follows.

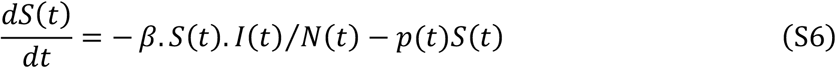

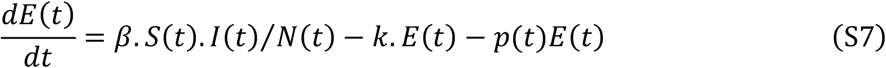

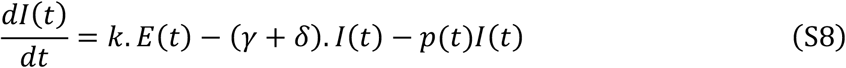

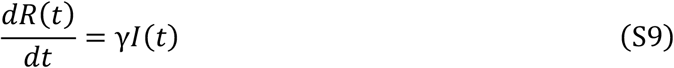

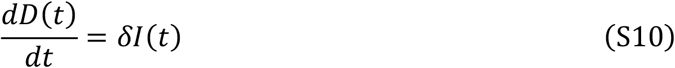

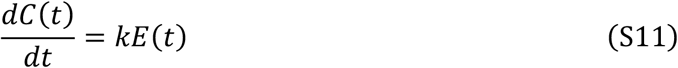

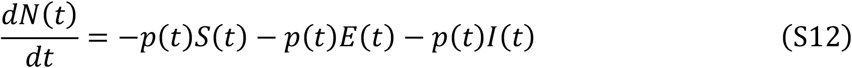

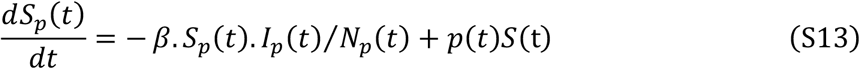

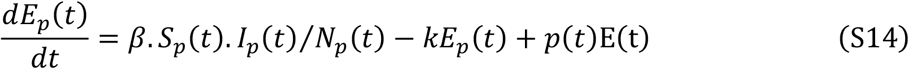

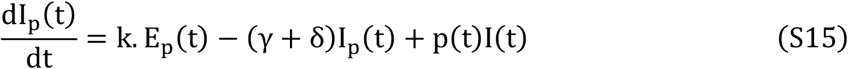

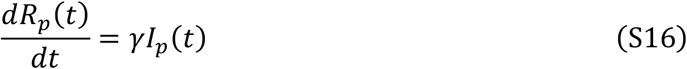

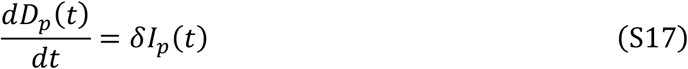

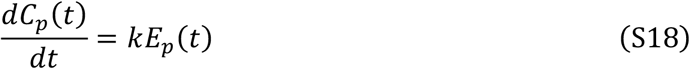

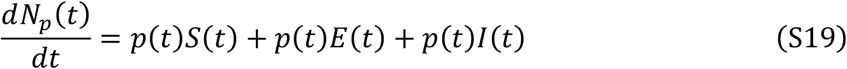

where *S*(*t*), *E*(*t*), *I*(*t*), *R*(*t*), and *D*(*t*) denote the number of susceptible, exposed, infectious, recovered and dead persons, respectively, in Wuhan. *N*(*t*) denotes the total population size. The variables with subscript p indicate that they are for the non-Wuhan area. Variable *p*(*t*) denotes the migration rate, the proportion of people moving from Wuhan to the non-Wuhan area. In the formulation, 1/*k* is the incubation period (7 days), 1/*γ* the infectious period (9 days), and *δ* = *γCPF*/(1 − *CPF*) is the death rate, which is estimated by the fatality rate 1.5%. These values were adopted from the WHO documents (*3*). We set December 4, 2019 as the starting date of the outbreak. By adopting the epidemiological characteristics reported in (*1*), we inferred the parameters for this customized SEIRDC model to fit our estimated number of L&I persons in Wuhan before the city’s lockdown. Following (*4*), the basic reproduction number *R*_0_ = 3.24 was obtained by fitting the growth of the confirmed case counts in mainland China excluding Hubei by January 22, 2020. The model-predicted epidemic size of 2019-nCoV is presented in Fig. 3 in the main manuscript.

**Table S1.** The number of people traveling from Wuhan (N), the cumulative number of confirmed cases (C), and the latent infection ratio (*l*) as of January 29, 2020.

| Province/Municipality | Number of people traveling from Wuhan (N) | Cumulative number of confirmed cases (C) | Latent infection ratio ($l = \frac{C}{N}$) |
| --- | --- | --- | --- |
| Wuhan | 9000000<br>(the number of people remaining in Wuhan) | 2261<br>(subject to under-screening and under-reporting biases) | — |
| Xiaogan, Hubei | 980896 | 399 | 0.41% |
| Huanggang, Hubei | 943987 | 496 | 0.53% |
| Jingzhou, Hubei | 474679 | 151 | 0.32% |
| Xianning, Hubei | 376580 | 130 | 0.35% |
| Ezhou, Hubei | 310037 | 123 | 0.40% |
| Xiangyang, Hubei | 287014 | 163 | 0.57% |
| Huangsi, Hubei | 281509 | 113 | 0.40% |
| Jingmen, Hubei | 232653 | 191 | 0.82% |
| Suizhou, Hubei | 225866 | 143 | 0.63% |
| Xiantao, Hubei | 215396 | 55 | 0.26% |
| Yichang, Hubei | 208748 | 117 | 0.56% |
| Tianmen, Hubei | 151180 | 44 | 0.29% |
| Enshi, Hubei | 134369 | 66 | 0.49% |
| Shiyan, Hubei | 131987 | 119 | 0.90% |
| Henan Province | 340467 | 278 | 0.82% |
| Hunan Province | 190635 | 277 | 1.45% |
| Guangdong Province | 103678 | 311 | 3% |
| Anhui Province | 99462 | 200 | 2.01% |
| Chongqing Municipality | 96420 | 165 | 1.71% |
| Jiangxi Province | 96081 | 162 | 1.69% |
| Beijing Municipality | 77854 | 111 | 1.43% |
| Shanghai Municipality | 60012 | 101 | 1.68% |
| Fujian Province | 33210 | 78 | 2.35% |
| Jiangsu Province | 38397 | 88 | 2.29% |
| Zhejiang Province | 44443 | 428 | 9.63% |
| Sichuan Province | 39621 | 142 | 3.58% |

**Table S2.** The cumulative number of confirmed cases as of January 29, 2020, with information concerning secondary transmission for the seven provinces and municipalities providing such information.

| Province/<br>Municipality | Cumulative<br>number of<br>confirmed<br>cases | Cumulative<br>Number of<br>secondary<br>transmission<br>cases | Cumulative<br>Number of cases<br>involving Wuhan<br>travelers | Proportion of non-secondary cases<br>(confirmed cases<br>involving Wuhan<br>travelers) |
| --- | --- | --- | --- | --- |
| Jiangsu | 88 | 7 | 81 | 92.05% |
| Beijing | 111 | 20 | 91 | 81.98% |
| Shanxi | 87 | 18 | 69 | 79.31% |
| Guizhou | 15 | 6 | 9 | 60.00% |
| Yunnan | 80 | 2 | 78 | 97.50% |
| Tianjing | 27 | 8 | 19 | 70.37% |
| Shanghai | 101 | 53 | 48 | 47.52% |

